# The Relatives’ Experience Questionnaire for Acute Inpatient Child and Adolescence Mental Health Services (REQ-AICAMHS): reliability and validity following a Norwegian survey

**DOI:** 10.64898/2026.05.06.26352577

**Authors:** Mona Haugum, Hilde Hestad Iversen, Kari Evelin Arellano Lorenzen, Johan Siqveland, Oyvind Bjertnaes

## Abstract

**Introduction:** Adolescents with mental health disorders represent a vulnerable group with complex care needs, yet their and their relatives’ experiences in acute inpatient mental health services remain poorly understood. While patient-reported experience measures (PREMs) are increasingly recognized as essential for improving healthcare quality, validated instruments for child and adolescent mental health inpatient settings are lacking—particularly from the perspective of relatives, who are often deeply involved in care.

**Objective:** This study aims to develop and psychometrically evaluate the Relatives’ Experience Questionnaire for Acute Inpatient Child and Adolescent Mental Health Services (REQ-AICAMHS), the first instrument designed to capture relatives’ experiences in Norwegian acute inpatient mental health units for adolescents.

**Methods:** The REQ-AICAMHS will be developed using the Norwegian Institute of Public Health’s (NIPH) standard methodology, including a literature review, expert consultation, qualitative interviews, and cognitive testing with relatives. Data will be collected digitally from relatives of all adolescents admitted to 17 acute inpatient child and adolescent mental health units across Norway. Descriptive statistics, exploratory factor analysis (EFA), and Item Response Theory (IRT) using the Generalized Partial Credit Model (GPCM) will be applied to assess data quality, factor structure, construct validity, and internal consistency. Open-ended responses will be analysed using both qualitative and machine learning methods to identify key themes and subdomains of experience.

**Results:** Preliminary results will include descriptive statistics of respondents, item-level analysis (missing data, ceiling effects), factor loadings and reliability coefficients (Cronbach’s alpha ≥0.7), inter-item and scale correlations, construct validity testing, and IRT parameters (item discrimination and threshold estimates). Findings will inform the development of a shortened version of the questionnaire for broader implementation in quality improvement initiatives.

## Introduction

Mental health disorders in adolescents present significant economic and public health concerns with lasting impacts [1], with over one in ten experiencing a mental illness at any given time [2]. Despite this, there remains limited knowledge regarding adolescent involvement in mental healthcare, and relatively few studies have assessed their experiences [1, 3, 4]. Patient experiences are acknowledged as vital to enhancing healthcare quality, alongside clinical effectiveness and patient safety [5-7]. A systematic review reported consistent positive links between patient experience, safety, and effectiveness across various conditions, care settings, outcomes, and study designs [5]. There is limited information about the routine application of patient-reported experience measures (PREMs) to enhance experience and outcomes in child and adolescent mental health services (CAMHS) [8].

Young people have distinct healthcare needs and perceptions, requiring tailored methods to measure their experiences [9]. A review identified eight key domains that shape young people’s care experiences: clinician attitudes, communication, competence, involvement, guideline-based care, outcomes, accessibility, and age-appropriate environments [10]. Within mental health services, adolescents particularly value participation in decision-making, privacy, age-relevant information, and a trusting relationship with professionals [8]. Consistency of staff is especially important in adolescent mental health care, as it supports trust and continuity; frequent staff turnover can disrupt therapeutic relationships and make it harder for adolescents to feel secure and supported [8].

Mental health issues among children and adolescents pose a major challenge in Norway [11]. The 2023–2033 White Paper emphasizes prevention, accessible services, and coordinated care for youths with complex needs [12]. Since 2015, discharges from child and adolescent inpatient units have increased [13], highlighting the urgent need for youth involvement in service development. Seventeen acute inpatient units serve this population, operating within the national quality network (Kvalitet i institusjonsbehandling i psykisk helsevern; KvIP [Quality of institutional treatment in mental health care]) at Akershus University Hospital and collaborating with service-user organizations on quality improvement [14, 15]. Despite peer visits and unit evaluations, systematic collection of individual user experiences remains uncoordinated and lacks validated instruments. Insights into accessibility, communication, coordination, and treatment experiences are essential to inform quality improvement efforts [16-18]. However, global reviews have found insufficient evidence on adolescent mental health service quality [19]. There is a lack of validated PREMs for mental health inpatients [20], making it impossible to assess service performance from the user perspective—diminishing potential for improved adherence and outcomes [6]. A recent doctoral thesis described the use of a user satisfaction questionnaire (the Experience of Service Questionnaire – ESQ) in outpatient CAMHS in Norway and the United Kingdom, however this questionnaire is not specifically developed for inpatients, and focuses primarily on satisfaction [21]. Although The Norwegian Institute of Public Health (NIPH) has developed outpatient PREMs [22, 23], none exist for acute inpatient settings in CAMHS, which involve unique experience domains like privacy, activities, and family engagement. Instruments tailored to capture these domains for youth and parents or caregivers – in this study named relatives – are therefore critically needed.

The NIPH conducts research on patient-reported experiences and outcomes, aiming to create a more patient-centred healthcare system through surveys and ongoing measurements across diverse patient groups [24-26]. Historically, the NIPH has concentrated on adult services, resulting in limited insight into the experiences of younger populations. Recently, however, new instruments have been developed to assess young patients’ experiences in outpatient diabetes care and CAMHS [22, 23]. The NIPH has also conducted national parent experience surveys in outpatient CAMHS in 2006 and 2017. The inclusion of both children and parents in a person-centred approach provides valuable insights into healthcare quality [27, 28].

Young people admitted to inpatient units are often accompanied by relatives, and sometimes the whole family. As part of a broader project developing PREMs for both adolescents and their relatives, given that no validated instruments currently exist for these groups specifically, the present study focuses on the relatives’ perspective. Specifically, it aims to evaluate the data quality, validity, and internal consistency reliability of the questionnaire for relatives accompanying adolescents admitted to mental health acute inpatient units. The questionnaire will be tested using standard methodology from the NIPH [24-26, 29-31]. The corresponding questionnaire for adolescents will be described in a second protocol and a separate publication.

## Material and methods

### Questionnaire development

The Relatives’ Experience Questionnaire for Acute Inpatient Child and Adolescence Mental Health Services (REQ-AICAMHS) will be developed and validated following the NIPH’s standard methodology. This includes a literature review, interviews with relatives, consultations with expert groups, and pilot testing [22, 23, 30, 31]. The purpose of the instrument is to evaluate relatives’ experiences during in-patient stays within a mental health acute inpatient unit.

A comprehensive literature review will be conducted to search for questionnaires, parts of questionnaires or topics that are considered important and relevant for the new questionnaire. An expert group will be consulted several times to discuss the content of the new questionnaire. The expert group will preferably consist of clinicians/therapists, researchers with extensive knowledge of the field, and representatives from patient organisations. Qualitative interviews will be conducted with relatives to make sure the questionnaire will be relevant, feasible and important. After summarizing the information gathered from the steps described above, we will construct the first version of the questionnaire, which we will cognitively test with a group of relatives. After this, some final adjustments will be made, and the questionnaire can be used in larger-scale-surveys in all the 17 mental health acute inpatient units.

Most questions in the questionnaire will be closed-ended; however, some open-ended questions will be included where respondents can write more about their experiences with the services, including improvement suggestions. This kind of data is highly valued by clinicians and well-suited for use in quality improvement [32]. For most PREMs items, the response categories are standardized to a five-point scale from 1 (not at all) to 5 (to a very large extent), with an additional “not applicable/don’t know” option. This response format is used consistently in NIPH surveys to allow for comparisons across time and user groups [25, 30, 31].

### Data Collection

The study has been judged as health services research, and falls therefore outside the Regional committees for medical and health research ethics’ mandate under the Health Research Act (P360: case number: 26/01457-1). The study was approved by the Institutional Review Board (IRB) located in the Health Services Division of the Norwegian Institute of Public Health (P360: case number: 26/01457-2).

For qualitative interviews, we plan to include 12-15 relatives. These respondents will be recruited with assistance from KvIP and the expert group. We will aim for the same number of respondents for the cognitive testing of the questionnaire. We will recruit adolescents and relatives at the same time, both to lighten the burden on those responsible for recruitment and to ensure that information from both groups can inform both questionnaires (the research protocol for adolescents will be published later). Informed consent is given verbally in the interview phases based on written information sheets and verbal information.

The relatives of all inpatients in the 17 mental health acute inpatient units in Norway will be eligible for inclusion. There are variations in size between the units, with the smallest having five beds and the largest having 23 (mean = 9.6) [33]. Six of the units have only acute beds, while the others also have elective or intermediate beds. We plan to include relative(s) of all patients, regardless of which type of bed they use. Mean number of yearly admissions for 15 of the 17 units were 123.87 (the lowest reporting 53 admissions, the highest reporting 208; two units did not report to KvIP this year), and the patients stayed on average for just over 11 days. The total number of admissions in 2025 for the 15 units who reported this were 1858 [33]. This variation in both unit size and patient flow warrant a longer and continuous data collection to ensure enough responses to produce valid and reliable results. The data can be collected either while the families are still at the units and close to discharge (preferred option to secure a high response rate), or soon after discharge. Given the relatively small gross sample size, particular attention will be required throughout the data collection process to secure sufficient responses and thereby ensure reliable and valid results.

Surveydata will be collected digitally. If collected after discharge, the participants will be identified using the Norwegian Patient Registry (NPR), with invitations distributed through Helsenorge.no, which reaches over 90% of Norway’s population. Everyone who is invited to participate in the pilot study receives an invitation letter about the study as a basis for informed consent, returning the completed questionnaire constitutes consent, which is the standard procedure in all patient-experience surveys conducted by the Norwegian Institute of Public Health. For children under 12, only parents will be invited to complete the questionnaire. Parental consent is mandatory for those under 16, while individuals aged 16 or older could consent independently. Guardians of 12–15-year-olds need to consent on behalf of the adolescent, making it sensible to collect data from the relatives first, and in a second wave, collect from the adolescents themselves. Non-responders will receive up to three SMS reminders. All collected data will be handled in compliance with the General Data Protection Regulation (GDPR) (EU 2016/679). Personal data will be anonymized where applicable, and appropriate technical and organizational measures implemented to ensure data security.

### Analysis

The responses to the open-ended questions will be analysed using both established qualitative methods and machine learning methods, both to validate the content of the instrument and to evaluate the whole comments (positive/negative) and subdomains in the comments (e.g.: information, communication, organization etc.).

Missing data and ceiling effects will be assessed. Items with more than 20% missing or “not applicable” responses will be excluded from factor analysis to reduce data loss [24, 30, 31]. Ceiling effects—defined as the percentage of respondents selecting the most favorable response—is acceptable if below 50% [24, 30, 31, 34].

We will conduct exploratory factor analysis (EFA) and apply Item Response Theory (IRT) to examine psychometric properties. For EFA, we principal-axis factoring with promax rotation will be used to identify the underlying factor structure of the REQ-AICAMHS. Factors with eigenvalues greater than 1 are retained. Items with factor loadings below 0.40 or cross-loadings above 0.30 are removed. The analysis will combine empirical and theoretical considerations, distinguishing between outcome-related items and those addressing structure and process. As recommended by Kilbourne et al., mental health quality measures should be validated using Donabedian’s model of structure, process, and outcome [35]. This model will inform our factor analysis, aiming to isolate distinct aspects of relatives’ experience.

Scale reliability will be evaluated using item-total correlations and Cronbach’s alpha, with 0.7 set as the minimum acceptable threshold [36]. To test discriminant validity, we will analyse how each item correlates with its hypothesized scale versus other scales.

Construct validity assesses how well patient-reported outcomes (HR-PRO) align with theoretical expectations, including internal correlations and differences between groups [37]. We will form hypotheses on how patient experiences and background factors will correlate.

To better understand item performance and support the development of a shortened instrument version, we will apply Item Response Theory and the Generalized Partial Credit Model (GPCM) [38]. This model estimates item discrimination and response category thresholds, focusing on structure and process items. Key parameters will include item discrimination (‘a’), item difficulty (‘b’), and the S–χ^2^ fit statistic. Higher ‘a’ values reflect greater sensitivity to the underlying trait; lower values suggest limited discriminatory power. S–χ^2^ indicates how well an item fits the model, with lower values suggesting better fit [39]. However, this statistic is sample-size sensitive, and large samples can show poor fit even when issues are minimal [38]. Item thresholds (b1 to b4) reveal at what point respondents are likely to endorse specific response levels. Higher negative thresholds indicate easier agreement; lower thresholds indicate greater difficulty.

The main goal is to create a concise version of the REQ-AICAMHS by selecting the most informative items based on missing data, ceiling effects, EFA, and IRT results. Agreement between the short and full “structure and process” scales will be assessed using intraclass correlation coefficients (ICCs). Analyses will be performed in SPSS and R, with specialized packages (lavaan, semPlot, mirt) for psychometric testing.

## Results

Table 1: Descriptive statistics of the respondents, e.g.: sex, age, background information

Table 2: Item description for the new questionnaire on relatives’ experience with mental health acuteinpatient units, e.g.: levels of missing data, mean values on items and ceiling effects

Table 3: Results from exploratory factor analysis, e.g.: factor structure, factor loadings, reliability statistics

Table 4: Results from correlations between items and the scales of the questionnaire

Table 5: Results from construct validity testing, i.e.: associations between scales, background variables and responses to questionnaire items

Table 6: Results from IRT analysis, i.e.: discrimination, difficulty thresholds, and item fit

### Availability of data and material

The datasets generated and/or analysed during the current study will not be publicly available due to the need to protect personal data but anonymous data are available from the corresponding author on reasonable request.

### Ethics

The study has been judged as health services research, and falls therefore outside the Regional committees for medical and health research ethics’ mandate under the Health Research Act (P360: case number: 26/01457-1).

The study was approved by the Institutional Review Board (IRB) located in the Health Services Division of the Norwegian Institute of Public Health (P360: case number: 26/01457-2).

The study will be performed in accordance with relevant guidelines and regulations, including the Declaration of Helsinki. Informed consent is given verbally in the interview phases based on written information sheets and verbal information. Everyone who is invited to participate in the pilot study receives an invitation letter about the study as a basis for informed consent, returning the completed questionnaire constitutes consent, which is the standard procedure in all patient-experience surveys conducted by the Norwegian Institute of Public Health.

